# A Statistical Argument Against Vaccine Injury

**DOI:** 10.1101/2022.04.19.22274036

**Authors:** Jacques Balayla

## Abstract

Vaccine hesitancy is a major threat to public health. While the root causes of vaccine hesitancy are numerous, they largely revolve around some form of perceived risk to the self. In particular, the unknown long-term risks are amongst the most frequently cited concerns. In this work, we show that regardless of their peak onset following vaccination, the incidence of adverse outcomes will follow some distribution *f* (*x*| *µ, σ*^2^) of mean onset *µ*, and standard deviation *σ*, and variance *σ*^2^. Despite the small proportion of events at the tails of these distributions, the large-scale public deployment of vaccines would imply that any signal for a given adverse outcome would be observed soon after distribution begins, even in cases where *t*_*x*_ *< t*_*µ−*3*σ*_. The absence of such an early signal, however low, would suggest that long term effects are unlikely and that vaccine safety is therefore likely. Indeed, when enough individuals have been exposed to a new therapy - even if the majority of adverse outcomes only manifest at a future time *t*_*µ*_, the number of adverse outcomes given by the cumulative density function (CDF) near *t*_0_ + *dt >* 0. Otherwise stated:

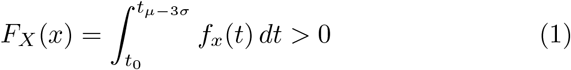

We evoke the theory behind normal (Gaussian) and skew-normal distributions and use Chebyshev’s Theorem to evaluate the COVID-19 vaccine data as an example. The findings of this study are not vaccine-specific and can be applied to assess the health effects of the mass distribution of any good, treatment or policy at large.

## 1 Background Theory

### 1.1 Gaussian Distribution and Central Limit Theorem

In mathematics, a *Gaussian function*, often simply referred to as the *Gaussian distribution curve*, is a continuous function of mean *µ* and standard deviation *σ* satisfying a number of properties herein described [1]. The Gaussian function is graphically represented by a bell-shaped probability density function (PDF) curve which approximates the exact binomial distribution [2]. The graphical representation of the curve is obtained by a function of the form:

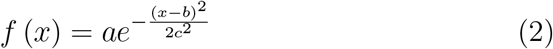

where a, b, and c are arbitrary real constants that satisfy the following properties: the parameter *a* reflects the height of the curve’s peak, b is the position of the center of the peak and c, the standard deviation, controls the width of the bell [3]. In probability theory, the Gaussian distribution is also known as the *normal* distribution, and its function is generalized to [4]:

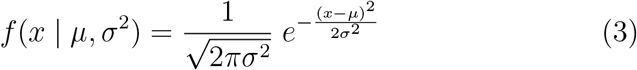

where: *µ* is the mean or expected value of the distribution, *σ* is the standard deviation and *σ*^2^ is the distribution’s variance [5]. The nature of the Gaussian function gives a probability of 0.682 of being within one standard deviation of the mean, a 0.954 probability of being within two standard deviations and a 0.997 probability of being within three standard deviations. The aforementioned statements are known as the 68-95-99 *empirical* rule, and can more easily de described as [6]:

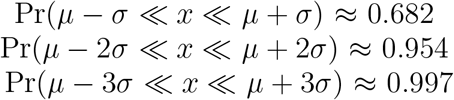

### 1.2 Central Limit Theorem

In its most general form, the Central Limit Theorem states that averages of samples of observations of random variables independently drawn from independent distributions converge in distribution to the normal, that is, they become normally distributed when the number of observations is sufficiently large [7]. With this theorem, we can transform every normally distributed bell-shaped function through standardisation into a new Gaussian function which has a mean of 0 and standard deviation of 1 [7]. The transformed data is denoted as the *standard normal*, where the probabilities are denoted by a Z-score. Simply put, a Z-score is a measure of numerical distance which translates into how many standard deviations below or above the population mean a raw score is found [7]. If some data *x* are normally distributed, the corresponding *z* will be normal with mean 0 and standard deviation 1 where the corresponding transformation between the *x* and *z* is given by the following standardisation formula [8]:

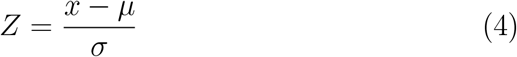

When *µ* = 0 and *σ* = 1 the generalized Gaussian equation in (2) becomes the *normal standard* function whose probabilities are denoted by Z scores. This special case of the Gaussian distribution is denoted ∼ *N* (0,1) and its equation is as follows [9]:

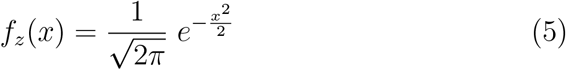

### 1.3 Basic Properties of Normally Distributed Variables

There are some interesting properties of normally distributed functions, which include the following: the mean, mode and median of the distribution are all equal [10]. The graphic representation of a normally distributed function is symmetric at the center, around the mean, *µ*. Consequently, exactly half of the values are to the left of center and exactly half the values are to the right [10]. The total area under the curve, which represents the sum of all probabilities is 1, as shown in the equation below:

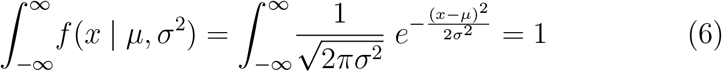

Note the presence of the mean, *µ*, which divides the curve symmetrically into different portions as a function of the standard deviation, *σ* (Figure 1):

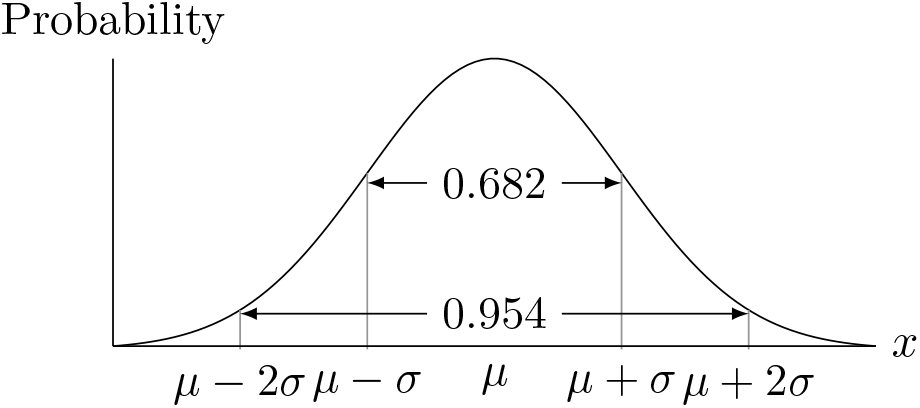

### 1.4 The Skew-Normal Distribution

Despite the fact a normal standard distribution is symmetric on either side of *µ*, such symmetry may not be appropriate to reflect the true distribution of a given set of data. Therefore, it is conceivable that some relationships might be better depicted by a different, *skewed* distribution. The skew normal distribution is a continuous probability distribution that generalises the normal distribution to allow for non-zero skewness [11]. A random variable *x* has a skew-normal distribution with parameter *α*, denoted by *x* ∼ SN(*α*), if its probability density function (PDF) is given by

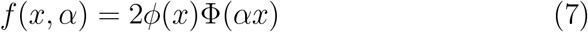

where Φ and *φ* are the standard normal cumulative distribution (SN-CDF) function and the standard normal probability density function (SN-PDF), respectively [12]. Herein, *x* and *α* are real numbers. Now, consider first a continuous random variable *φ*(*x*) denoting the standard normal Gaussian probability density function (PDF), exhibiting a mean *µ* = 0 and *σ* = 1, as in equation (5):

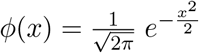

Its corresponding CDF, Φ(*x*), using a dummy variable *t* is:

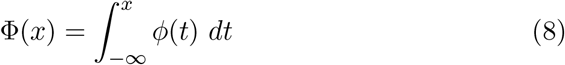

In the presence of a skewness coefficient *α*, also known as the *shape parameter*, Φ(*x*) becomes Φ(*αx*):

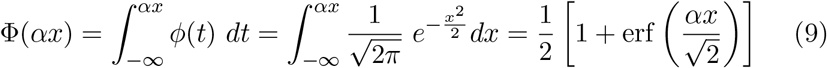

The **erf** stands for the Gauss error function [13]. In mathematics, the Gauss error function is a special and non-elementary function of sigmoid shape that occurs in probability, statistics, and partial differential equations describing diffusion. It is defined by the following equation[13]:

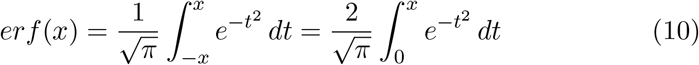

The integral in equation (10) can be expanded and approximated through a McLaurin series (Taylor Series with parameter *a* = 0) as follows [14]:

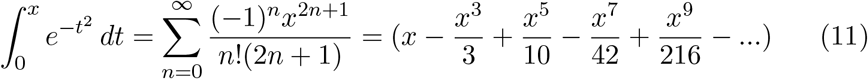

Since the series is uniformly convergent, it may be integrated term by term. By replacing *x* for the interval in question, Φ(*αx*) becomes:

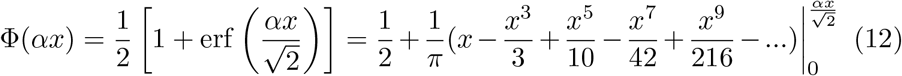

In its complete form, the skew-normal distribution requires location, scale and shape parameters. These are real numbers and defined by *ξ, ω* and *α*, respectively, so that:

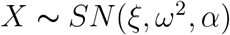

With these parameters, the probability density function (PDF) of the skew normal distribution in equation (7) is generalized to:

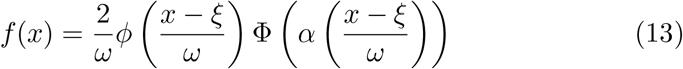

which can otherwise be written as follows:

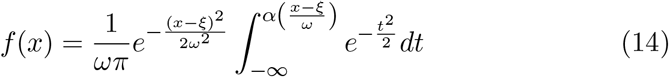

### 1.5 Basic Properties of the Skew-Normal Distributions

The probability density function of the skew-normal distribution enjoys various interesting formal properties. First, when *α* = 0, the skewness vanishes, and we obtain the *standard normal* density function. Secondly, as *α* increases, the skewness of the distribution increases as well. Thirdly, as *α* → ∞, the density converges to the so-called half-normal (or folded normal) density function. And lastly, if the sign of *α* changes, the density is reflected on the opposite side of the vertical axis. The characteristics of a skew normal distribution curve are a function of its parameters. For simplicity’s sake, we first define *δ* as follows:

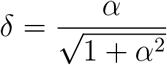

The mean *µ*_*k*_ can thus be described by the following relationship:

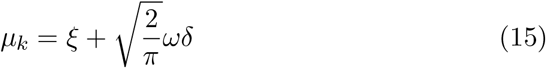

The variance Var is given by:

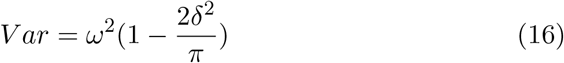

The skewness *γ* is defined as:

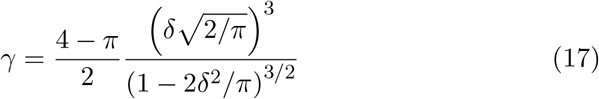

The kurtosis *κ* is given by:

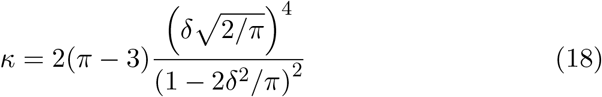

### 1.6 Location - Xi (*ξ*)

The location parameter *ξ* is a scalar- or vector-valued parameter, which determines the degree of shift of the distribution in question relative to a standard. For time dependent distributions, higher values of *ξ* depict probabilities that arise later in time and vice-versa. For non-time dependent distributions, the parameter *ξ* remains largely inconsequential, since it does not alter the shape nor the relative distribution of probabilities.

### 1.7 Scale - Omega (*ω*)

The scale parameter *ω* is a scalar, which depicts the statistical dispersion of the probability density function. The larger the scale parameter, the more spread out the distribution, and vice versa.

### 1.8 Shape - Alpha (*α*)

The shape parameter *α* describes any and all parameters that do not describe scale or the location of the density function. As such, *α* is directly related to the derivative of the probability density function.

### 1.9 Examples of Skew-Normal Distributions

**Figure 2.**
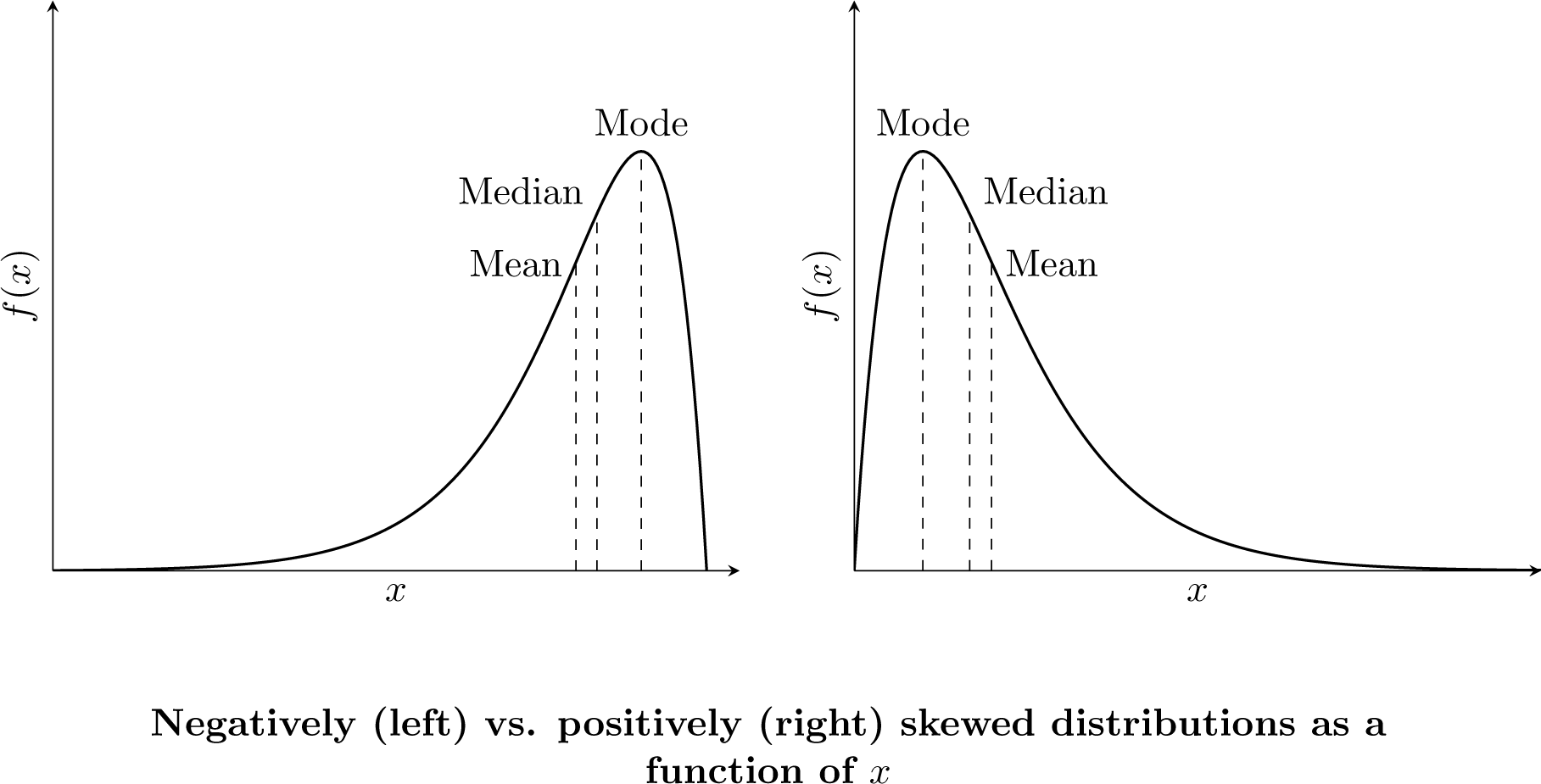
Random positively and negatively skewed distributions

Unlike the normal distribution, the skew-normal distribution has different mean, mode, and median values which different from one another. The variations between different skews can be significant and in particular, differences in *α, ω* and *ξ* render skew distributions quite versatile and amenable to describe a number of phenomena in the natural world. These distributions can portray a cross-sectional picture at any one time, e.g. the height of students in a classroom or simply reflect the onset probability of some phenomenon *x* over some time *t*, as we will see in the following section.

## 2 Applications of the theory in vaccine programs

To preface this section, let us come up with a hypothetical scenario:

*The year is 2019. A novel virus with great contagious potential has been detected. The spread becomes inevitable and soon after a pandemic ensues. Thousands of people die every day worldwide. No clear therapeutic agent is identified. However, in an inspiring international effort, a vaccine is developed using novel technology, and it is soon after distributed all over the world. To date, 10*.*86 billion vaccines have been administered - and 63*.*5*% *of the world population has received at least one dose. Despite the critical threat the virus poses - vaccine hesitancy becomes a prominent force in societies. The reasons are multiple, but they all revolve around some form of perceived risk to the self. In particular, the unknown long-term risks are amongst the most frequently cited concerns*.

Are these concerns founded? If so, are these concerns time-dependent? In other words, how long after the vaccine roll-out do concerns about injury become unfounded? We hereby provide a statistical argument against vaccine injury regardless of the nature of the risks in question - as their nature is irrelevant to the argument presented here.

Let *x* represent the incidence of some adverse event following vaccination. Let *f* (*x*) represent its probability density function over some time *t*. Because we don’t know what kind of distribution *f* (*x* | *µ, σ*^2^) has, we consider two distinct scenarios: one where the *x* follows the normal distribution, and one where it follows a skewed one.

### 2.1 Model 1: f(x) follows a normal distribution

If *x* follows a normal distribution with mean *µ* and standard deviation *σ*, then as per equation (3), three standard deviations (3*σ*), would cover 99.7% of the area under the probability density function, which is the equivalent of the cumulative distributive function [6]. Because the distribution is twotailed, the area below −3*σ* needs to be divided by 2. Let us for arguments’ sake define the time of vaccine roll-out as *t*_0_ and the time of mean onset of adverse outcomes as *t*_*µ*_ so that *t*_0_ *< t*_*µ*_. The left “tail” of the distribution curve begins at *t*_0_ such that *t*_0_ *< t*_*x*_ *< t*_*µ−*3*σ*_ [6]. Despite the rather small proportion of all adverse outcomes that lie at such extremes of the distribution (0.003%), the sheer number of vaccines administered would yield a signal at the onset of the public deployment. As time evolves, such a signal would gain intensity such that the peak number of cases is observed at *t*_*µ*_.

**Figure 3.**
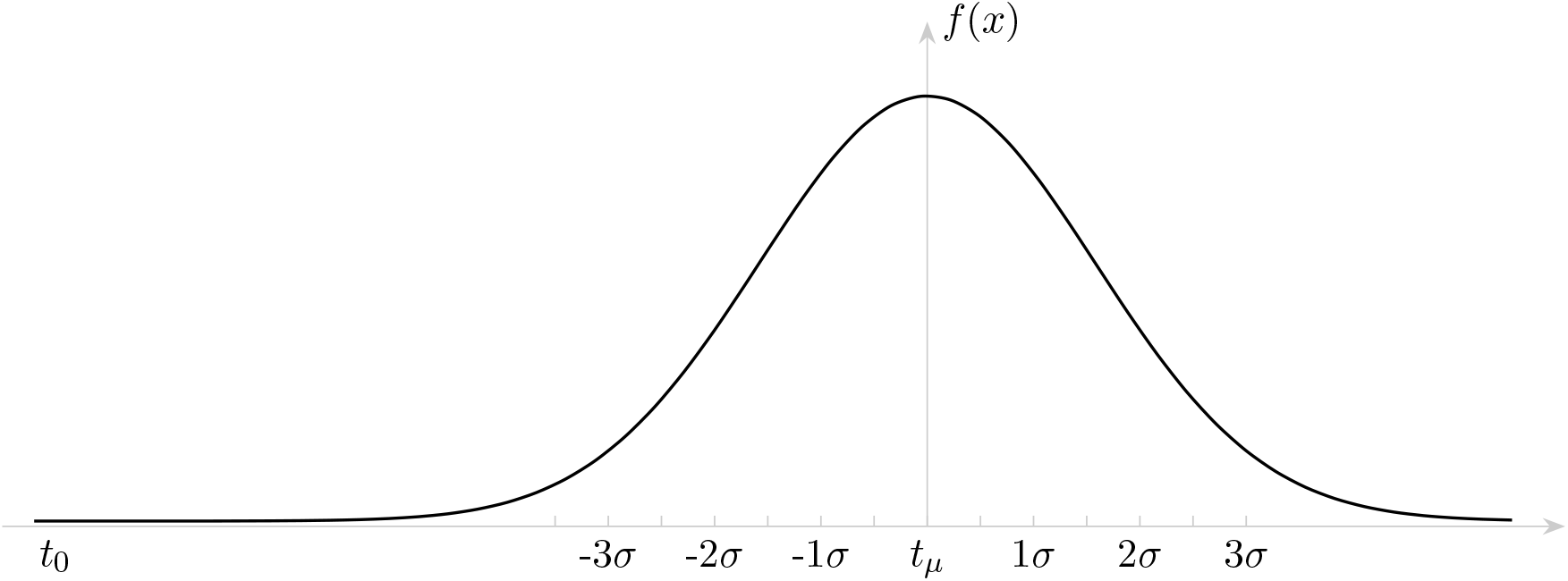
Sample normal distribution of the incidence of adverse outcomes beginning at *t*_0_

While the above figure appears to suggest that below −3*σ* from *t*_*µ*_, *f* (*x*) = 0, this is not so. Indeed, zooming in on the left “tail” we would observe that the CDF function at that point is non-zero.

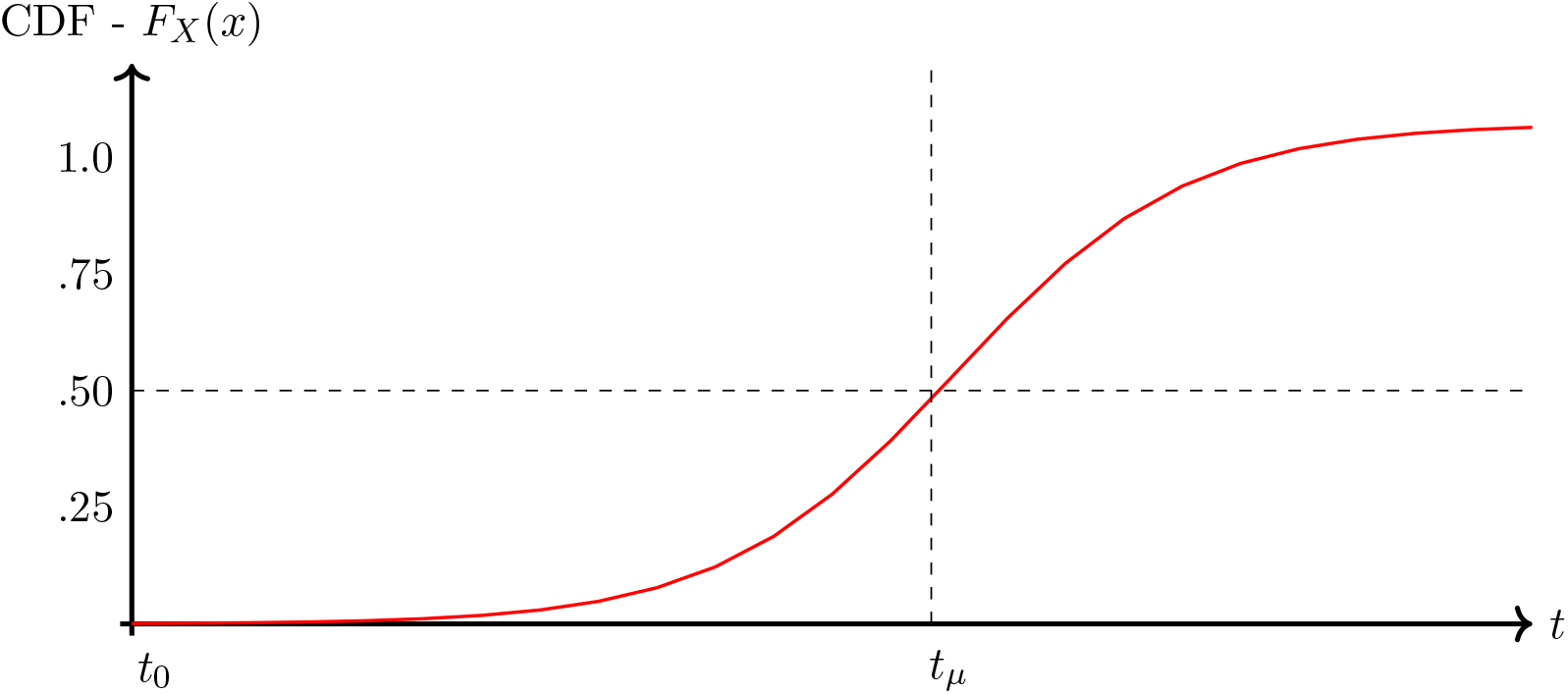

As is readily observed, the CDF(x) is therefore non-zero beyond *t*_0_ such that:

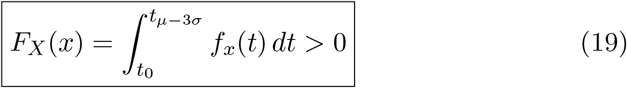

If we take the COVID-19 outcome as an example of a pandemic for which a vaccine was developed, we retrieve 11.4B vaccines given over the last 2 years [15]. Even if we assumed a very rare rate of of vaccine injury of say, 1% of 1% (0.0001) - implying 99.99% safety - at levels below −3*σ* (Table 1), we would obtain the following number of adverse outcomes directly related to the vaccine:

**Table 1.**
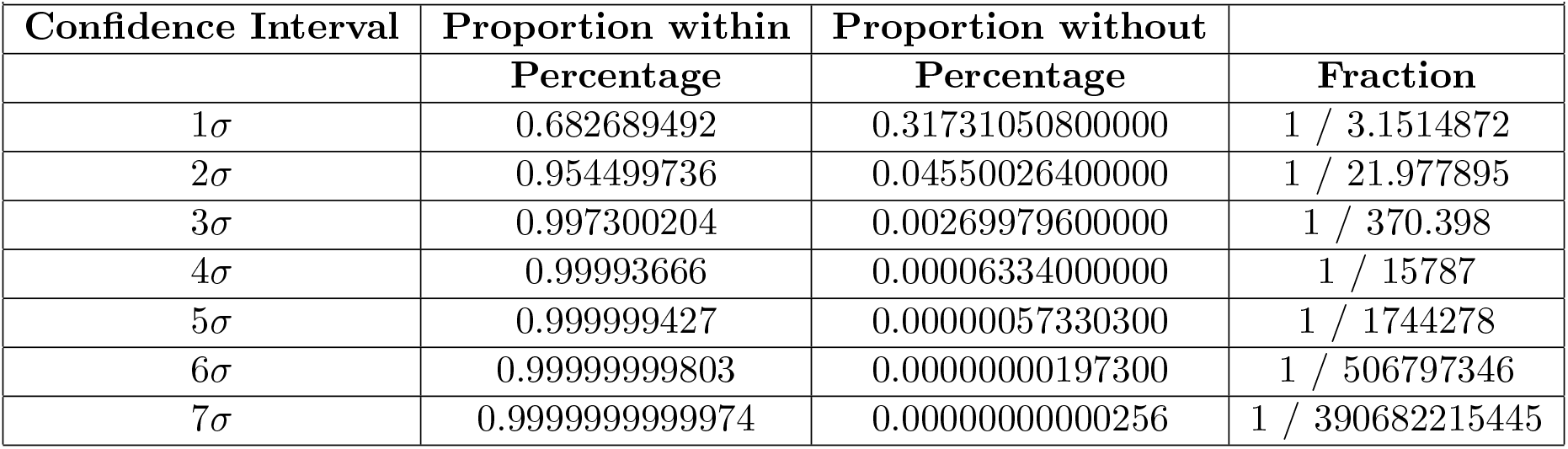
Proportion of the AUC of the PDF as a function of *σ*

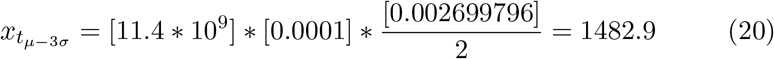

While this number appears low, it represents only 0.0013% of all adverse outcomes. Though this signal would appear early following mass distribution, it is only one of the possible adverse outcomes to be considered. Should such a signal not be observed, it is thus statistically unlikely that adverse outcomes directly related to the vaccine will occur significantly at any point at some future time *t*. On the other hand, the virus itself may very well be lethal - killing significantly more people in a day than the theoretical calculation above so the risk calculation ought to account for the lethality of the virus itself as well. Indeed, if 1482 individuals experience adverse outcomes related to the vaccine before −3*σ* at a rate of 1% of 1% (0.0001) which represents 0.002699796/2% of all outcomes, then we would expect when *t > µ* + 3*σ* the following number of adverse outcomes related to the vaccine:

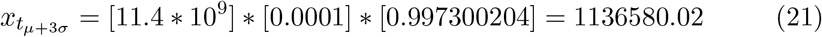

Note that the rate of injury here is not divided by two as the symmetry of the Gaussian implies that the outcomes on the left tail have already happened and are thus accounted for in the calculation. Since the total number of COVID deaths is about 6.19 million worldwide to date - this would imply that the rate of vaccine injury amount to 1/6 of all deaths - a staggering number! Once again, the absence of the aforementioned early signal would suggest vaccine injury is unlikely.

### 2.2 Criticisms of this model

It is conceivable that if *µ* is sufficiently far away, that even two years into the pandemic, we may not have attained the 3*σ* level below which you only should see 0.002% of adverse outcomes yet. In other words - that we’re still too early to detect the long-term adverse outcomes. However, it bears saying, there’s no historic comparison where adverse outcomes of a new vaccine are seen years down the road nor is there any physiological basis from which to suspect such long-term vaccine injury. After all - the reasons we have needed the boosters is because the immunity provided by the vaccine weans over a time frame of months - not years [16]. That we have not already directly observed deaths or adverse outcomes beyond that which is expected should re-assure individuals as to the absence of long-term risk following vaccination. As alluded to above, this is not COVID-specific and can be applied to any vaccine or mass-distributed product with potential health implications.

### 2.3 Model 2: f(x) follows a positively-skewed distribution

If we consider the same time axis *t* but instead take on a skew-distribution of *f* (*x*) we obtain two possibilities: a positively-skewed distribution and and negatively-skewed distribution. The case of the positively-skewed distribution is straight forward as the mean is close to *t*_0_, and thus the onset of any adverse outcome would appear in significant numbers close to *t*_0_. In other words, any signal of adverse outcome - should it exist - would be amplified early on and peoples’ apprehensions would either be confirmed or denied soon after mass deployment of the vaccine.

### 2.4 Model 3: f(x) follows a negatively-skewed distribution

If we consider the negatively-skewed distribution, it could be plausible that the distribution’s peak occurs further into the future than in the case of a normal distribution. Unfortunately, we cannot always strictly speak of standard deviation in a skewed-distribution and may instead opt to speak of interquartile ranges (Q1, Q2…) often using boxplot-whiskers as representations of skewed distributions [17]. Though the shape of the curve is different we can still use the CDF function to determine the absolute number of adverse outcomes observed at some time *t* following vaccine distribution.

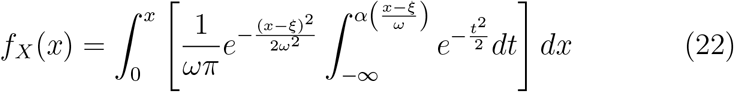

Is this integral always positive? It would stand to reason that if the skew-normal distribution is always positive, the area under its curve is positive as well. To prove this, we equate equation (14) to zero:

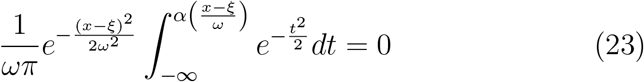

Dividing both sides by the integral term we obtain:

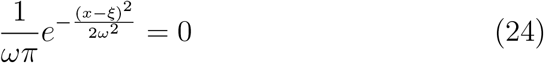

Dividing both sides by the constant term 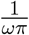 we obtain:

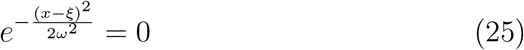

It is thus clear that 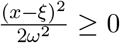 and as such:

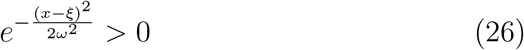

Another way to prove the above statement is to take the natural logarithm of either side and realizing that ln(0) is undefined, hence the relationship in (25) cannot hold. It thus follows that the skew-normal distribution always holds positive values and as such, its corresponding cumulative distributive function (CDF), which evaluates the AUC at some point *x*, is always positive as well. We thus conclude that:

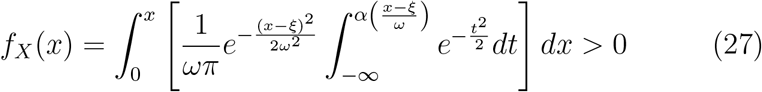

for all values of x.

## 3 Chebyshev’s Theorem

Chebyshev’s theorem, also known as Chebyshev’s inequality, suggests that regardless of the shape of a distribution one can determine the minimal and maximal bounds of the data included within *k* standard deviations from the mean *µ*. In technical terms, the theorem suggests: “For any numerical data set, at least 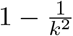of the data lie within *k* standard deviations of the mean, that is, in the interval with endpoints *x* ± *ks* for samples and with endpoints *µ* ±*kσ* for populations, where k is any positive whole number that is greater than 1”. Since *k* is by definition *>* 1, a positive proportion of the data lies even at remove values of *k* suggesting that the CDF, as above, will be non-zero. Again, with sufficient mass distribution of vaccines, the signal for early adverse outcomes would be manifest and would inform providers and the public as to the risks of vaccination.

## 4 Conclusion

As we have thus proven in this manuscript, the onset of adverse outcomes will follow some distribution following vaccine deployment. Whether the distribution is normal or skewed, the cumulative density function (CDF) will always be positive, and as such, any adverse outcome seen in significant levels would be observed soon after vaccines are deployed. Absence of such a signal should be interpreted as a strong argument against vaccine injury and ought to re-assure both vaccine providers and recipients as to the safety of the vaccine in question.

## Data Availability

No data was utilized in this study

